# Seizure prophylaxis in glioma surgery (SPRING): a multi-centre, unblinded, randomised trial

**DOI:** 10.1101/2025.09.03.25335027

**Authors:** Michael D Jenkinson, Helen Bulbeck, Jackie Burns, Alasdair G Rooney, Richard Dobbie, Jade Carruthers, Sarah Lessels, Rachael Watson, Ciara Gribben, Tomos Robinson, Luke Vale, Gerard Thompson, Sara C Erridge, Colin Watts, Anthony G Marson, Robin Grant

## Abstract

**Background:** 50-80% of all glioma patients will have seizures. Guidelines recommend against prophylactic anti-seizure medication, but levetiracetam is frequently prescribed. Our aim was to determine the effectiveness of 12-months prophylactic levetiracetam at reducing seizure risk.

**Methods:** Seizure-naïve patients undergoing glioma surgery were randomised (1:1) to 12-months levetiracetam or no prophylaxis and followed until death or maximum 18-months. The primary outcome was one-year risk of first seizure. Patients who died within 12-months of randomisation without experiencing a seizure were excluded from the primary outcome analysis. Target accrual was 804 participants. The trial was registered (ISRCTN 49474281 and EudraCT 2018-001312-30).

**Results:** Between Oct 10, 2019 and Aug 30, 2022, 96 patients, from 24 to 79 years of age, were randomised to levetiracetam (n=49) or no prophylaxis (n=47). The trial closed early due to slow accrual and the Covid pandemic. In the levetiracetam group 17 patients had a seizure and 32 did not (19 survived ≥ 12-months, 13 died within 12-months). In the no prophylaxis group 15 patients had a seizure and 30 did not (21 survived ≥12-months, 9 died within 12-months). Seventeen of the 36 evaluable levetiracetam patients (47%) had a seizure, compared with 15 of 36 evaluable no prophylaxis patients (41%) (OR 1.25, 95%CI 0.49-3.21, p=0.64). There were no levetiracetam related serious adverse events.

**Conclusions:** The SPRING trial provides no evidence of a difference between levetiracetam and no prophylaxis in the 12-month seizure risk in patients undergoing glioma surgery, but the study was underpowered. The role of prophylactic anti-seizure medication remains undefined.

## Introduction

In patients who have a glioma, 20-25% will present with a new onset seizure. Of the remaining 75-80%, one-third to a half will develop seizures at some point during treatment or at a later stage prior to death.(5 13 25) In the majority of cases infiltrative glioma are not curable and consequently optimising symptom control is important for both patients and carers. Seizures have a major impact on quality of life and may result in injuries or life-threatening complications such as status epilepticus or aspiration pneumonia.(2) Patients may also suffer debilitating anxiety about whether or when a seizure may occur.(2)

Anti-seizure medication (ASM) are administered to those with seizures, but there remains controversy regarding the benefit of prophylactic ASM to seizure-naïve patients with glioma.(24) The ability to prevent seizures is of great importance to patients and clinicians. There has been a shift away from the use of first generation drugs such as phenytoin, to newer generation ASM such as levetiracetam.(8) Guidelines from Society for Neuro-Oncology (SNO) and European Association for Neuro-Oncology (EANO) state that there is insufficient evidence to support prescribing prophylactic ASM at the point of diagnosis or the time of surgery, but these guidelines are based on trials in all brain tumors using older drugs such as phenytoin and valproate,(2 26) meaning the evidence is both dated and not directly relevant to this clinical context. Despite this recommendation, prophylactic ASM continues to be prescribed pre-operatively.(8) Levetiracetam is the most frequently used ASM in patients with glioma since it is a non-enzyme inducing so does not interact with dexamethasone, proton pump inhibitors, or chemotherapy. Levetiracetam has a lower side effect profile compared to older drugs, but still causes fatigue and behavioural problems, which may be compounded by the symptoms from the glioma, surgery and oncological treatment.(2) Although short course peri-operative use is most common(15), there are observational studies suggesting that the duration of treatment prophylaxis is important and those who receive prophylaxis for 6-months or more had significantly fewer seizures than those on prophylaxis for less than one-month or not at all.(18) This may be due to the higher risk of seizures at glioma relapse or prior to death. A systematic review recommended future research should focus on the role of prophylactic anti-seizure medication in the end of life phase, particularly in high grade glioma patients whose survival is often measure in months(17) and that clinical trials should use newer ASM such as levetiracetam.(24 26)

The Seizure Prophylaxis In Glioma multi-center randomised controlled trial (SPRING) was conducted to assess the clinical effectiveness of 12-months levetiracetam at reducing the risk of seizures, compared to no anti-seizure medication in seizure-naïve patients undergoing surgery for their glioma.

## Methods

### Study design

In this multi-centre, unblinded, randomised controlled trial 12-months of prophylactic levetiracetam was compared to no anti-seizure medication in seizure-naïve patients undergoing surgery for a newly-diagnosed glioma. Trial sites were 14 regional adult neurosurgery and neuro-oncology centers in the United Kingdom and Eire (See Section 1 of the Supplementary Appendix). Ethics approval was obtained from the East of England – Essex Research Ethics Committee (ref: 18/EE/0389). The trial protocol is available at https://fundingawards.nihr.ac.uk/award/16/31/136 (Substantial amendments are detailed in Section 6 of the Supplementary Appendix).

### Participants

To undergo randomisation in the trial, patients had a cerebral glioma on MRI and planned for surgery (biopsy or resection), were ≥ 16 years old, and had a Karnofsky Performance Status (KPS) ≥70. Patients were excluded if they were pregnant, had a seizure in the 10 years prior to randomisation, severe chronic kidney disease (CKD4 - eGFR < 30ml/min), were taking concomitant methotrexate, other anti-seizure medication or benzodiazepines, had hypersensitivity to levetiracetam, or had active suicidal ideation or severe depression as defined by a PHQ-9 score of ≥ 20. Patients gave written informed consent.

### Randomisation and masking

Patients were randomized to 12-months prophylactic levetiracetam or no anti-seizure medication at a ratio of 1:1 using a minimization algorithm. The randomization sequence was stratified by neurosurgical unit, glioma grade (high versus low) and surgery type (biopsy or resection). Randomization occurred prior to surgery using the Edinburgh Clinical Trials Unit (ECTU) randomization system, a web-based randomization tool. Patients and clinicians were not blinded to the allocation or the assessment of outcomes.

### Procedures

Levetiracetam was taken orally in two daily doses of 500mg approximately 12 hours apart. Patients with impaired renal function (CKD 3, eGFR 30-59 ml/min/1.73m^2^) started on a lower dose (25omg bd) for the first 2 weeks and then titrated up (Table S1). All patients received a minimum of 2 doses prior to surgery and from 2-weeks to 12 months were taking 750mg bd (or 500mg bd in patients with impaired renal function). After completing 12-months of levetiracetam, patients who did not have a seizure were placed on a 6-week tapering regime to discontinue the drug (Table S1).

Data were collected at pre-surgery (baseline), early post-operative assessment; and 3-monthly thereafter (Table S2). Patients were followed for a minimum 12-months and maximum 18-months. The types of data collected at each visit and the methods used are detailed in the online study protocol.

### Outcomes

The primary objective was to determine in seizure-naive newly diagnosed cerebral glioma patients undergoing surgery, whether prophylactic levetiracetam pre-operatively and for at least 1 year, produces a meaningful (>50%) reduction in the risk of developing seizures, when compared with standard care (no ASM). The corresponding primary outcome was 12-month risk of first seizure. Seizures were defined as simple partial, complex partial or partial seizure with secondary generalisation (see panel). Seizures were assessed locally by the treating clinical team. In cases of diagnostic uncertainty, patients were reviewed by neurologists as part of the routine clinical pathway to confirm or refute the diagnosis. Excluded attacks were those deemed by the treating clinician not to be epileptic seizures.

#### Panel: seizure definitions

1. Focal aware seizures (formerly ‘simple partial seizures’):

a. with motor symptoms: focal motor movements, versive/postural movements
b. with sensory symptoms: olfactory sensations
c. with autonomic signs
d. with psychic symptoms (e.g. déjà vu, jamais vu)
2. Focal seizures with altered awareness (formerly ‘complex partial seizures’):

a. with impairment of consciousness only
b. with impairment of consciousness plus automatisms (lip smacking, fumbling, etc.)
3. Focal to bilateral tonic clonic seizures (formerly ‘partial seizures with secondarily generalized seizures’):

a. Unconsciousness with generalised clonic movements
b. Unconsciousness with generalised tonic spasm, without clonic movements
c. Unconsciousness or staring with one of the following symptoms perceived by the patient:

- A rising feeling from the abdomen to the throat
- Smelling of odd scents
- Stiffening or convulsions in the face or limb(s)
- Turning the head to one side

Secondary outcomes were: time to first seizure; time to first tonic clonic seizure; patient reported symptoms and adverse events; mood, personality, fatigue and memory; progression free survival (PFS) and overall survival (OS). Health economic outcomes were EQ-5D-5L quality-adjusted life years (QALYs) at 12-months, and costs to the NHS and personal social services. A safety analysis was performed only for those patients randomized to levetiracetam. Data on serious adverse events were collected (see online protocol).

### Statistical analysis

A Trial Steering Committee, comprising a majority of independent members viewing reports blinded to treatment arm, and an Independent Data Monitoring Committee viewing unblinded reports reviewed the trial regularly to assess conduct, progress including rates of seizures, and safety. The sample size for the primary outcome was calculated based on a 20% 12-month seizure rate in patients with suspected cerebral glioma after surgery. The trial hypothesis was a reduction in seizure rate to 10% in the treatment arm. Based on a 90% power to identify an improvement in 12-month seizure rate in the treatment arm compared to the control arm, required 532 patients across the two arms with a two-sided type I error level of 5%. Assuming a 24.8% 12-month mortality rate and that 12% of patients will be lost to follow-up, the final (maximum) sample size was 402 per arm.

The analysis was conducted according to a prespecified statistical analysis plan. Outcomes were analysed according to the intention to treat principle with a 5% level of statistical significance and 95% confidence intervals. The occurrence of seizures within 12-months of randomisation was compared between study arms in those patients who survived for 12 months, using a logistic regression model fitted to the presence of a seizure within 12-months of randomisation. The odds ratio (OR), associated 95% confidence intervals and 2-sided p-values associated with the comparison of each arm from are given. The estimated absolute difference in seizure rate is reported together with associated 95% confidence interval. The estimated absolute difference in seizure rate is reported together with the associated 95% confidence interval.

For secondary outcomes all patients were included, and the mean and median time to first seizure and time to first tonic-clonic seizure was calculated for each study arm, and the seizure free probability within 12 months was analysed using Kaplan-Meier plots. Progression free survival (PFS) and overall survival (OS) were also analysed using Kaplan-Meier plots. The time to event distributions are statistically compared using log-rank tests based on the chi-square distribution with one degree of freedom. Due to the small number of events, the impact of covariates on time to event were not modelled using Cox Proportional Hazards. Quality of life, semantic verbal fluency test, anterograde memory and PHQ-9 scores were analysed and presented for the two randomisation groups but no statistical comparisons were made due to the small patient numbers.

Primary outcome and safety analyses were validated by independent programming from the point of raw data extraction. All analyses were done using R packages in a Posit Workbench. The trial was registered with ISCTRN: 70051203 and EudraCT 2018-001312-30.

### Economic analysis

The economic analysis adopted the perspective of the National Health Service (NHS) and personal and social care services in the UK. As the SPRING trial was closed prior to reaching the recruitment target, the planned within-trial analysis and long-term economic model were considered to no longer be appropriate. Instead HRQoL and health service utilisation data from the intention to treat population were described using summary statistics but no formal statistical comparisons were made between the two arms.

The responses to the EQ-5D-5L were converted to utility scores using the mapping function developed by the NICE Decision Support Unit (DSU),(12) using the ‘EEPRU dataset’.(11) Data regarding the dispensing of the intervention drug for each participant in the intervention were calculated using information from the dosage and were micro-costed. The price of Levetiracetam was gathered from the British National Formulary (BNF).(1) Data on health service resource use of the trial participants in the two trial arms were collected using a health service utilisation (HSU) questionnaire administered to all participants at 3-months, 6-months, 9-months and 12-months post baseline. Additionally, a Time and Travel (T&T) questionnaire was completed at a single time point (6-months). Unit costs were gathered from the Unit Costs of Health and Social Care and National Schedule of NHS Costs. The cost year of the analysis was 2023. The results from a standard gamble sub-study have already been reported.(21)

## Results

Between October 9, 2019 and August 31, 2022, 107 patients were recruited of which 13 were ineligible for randomisation. There were 94 patients randomly assigned to the study arms (49 to levetiracetam, and 45 to no prophylaxis; Figure 1). The characteristics of the two groups were similar at baseline (Table 1) and balanced for Karnofsky Performance Status (Table S3). All recruited patients had a glioma diagnosis and the anatomical location, histopathology diagnosis and molecular features are shown in Tables S4-S6.

**Figure 1:**
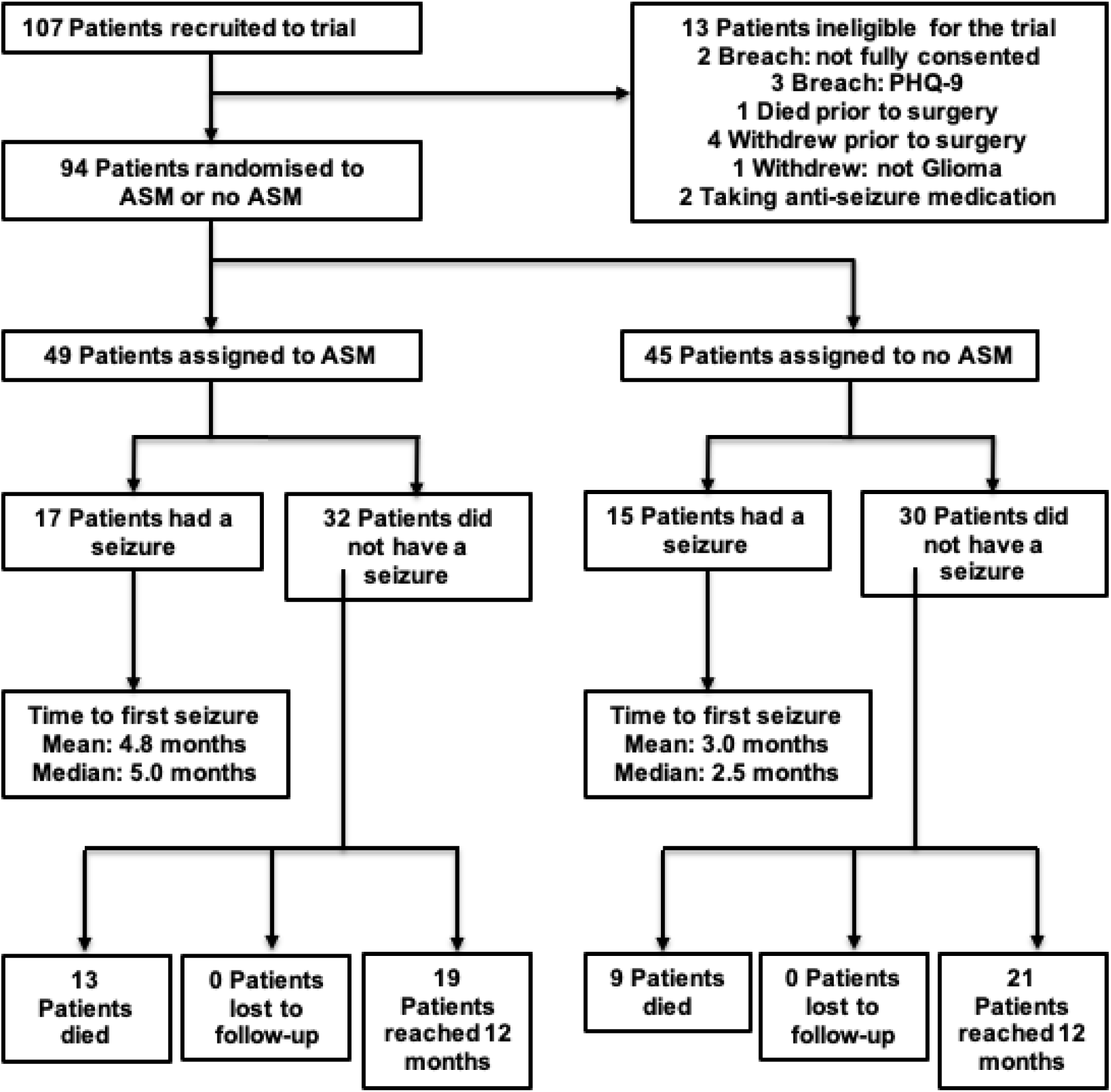
Trial profile. ASM; anti-seizure medication.

**Table 1:** Baseline characteristics of the intention to treat population.

|  | **Intervention arm**<br>**(Levetiracetam)** | **Control arm**<br>**(no prophylaxis)** |
| --- | --- | --- |
| **Age (years)** |  |  |
| Median | 61 | 59 |
| Range | 24-79 | 27-78 |
| **Sex** |  |  |
| Female | 15 | 18 |
| Male | 34 | 27 |
| **Glioma Grade (based on MRI)** |  |  |
| High grade glioma | 43 | 41 |
| Low grade glioma | 6 | 4 |
| **Surgery** |  |  |
| Biopsy | 7 | 6 |
| Resection | 42 | 39 |

Patients who died within 12-months of randomisation without experiencing a seizure (n=13 in the levetiracetam group and n=9 in the no prophylaxis group) were excluded from the primary outcome analysis. Compared to the no-prophylaxis group (15 out of 36; 41%) the levetiracetam group (17 out of 36; 47%) had an increased likelihood of seizures (OR 1.25, 95% CI: 0.49–3.21, p=0.64) but the difference was not statistically significant. In patients who developed seizures, the median time to first seizure was 5.0-months in the levetiracetam group and 2.5-months in the no prophylaxis group. For the whole study population the 12-month seizure free rate was 58.8% in the levetiracetam group and 63.8% in the no prophylaxis group and there was no statistically significant difference between the groups (Figure 2).

**Figure 2.**
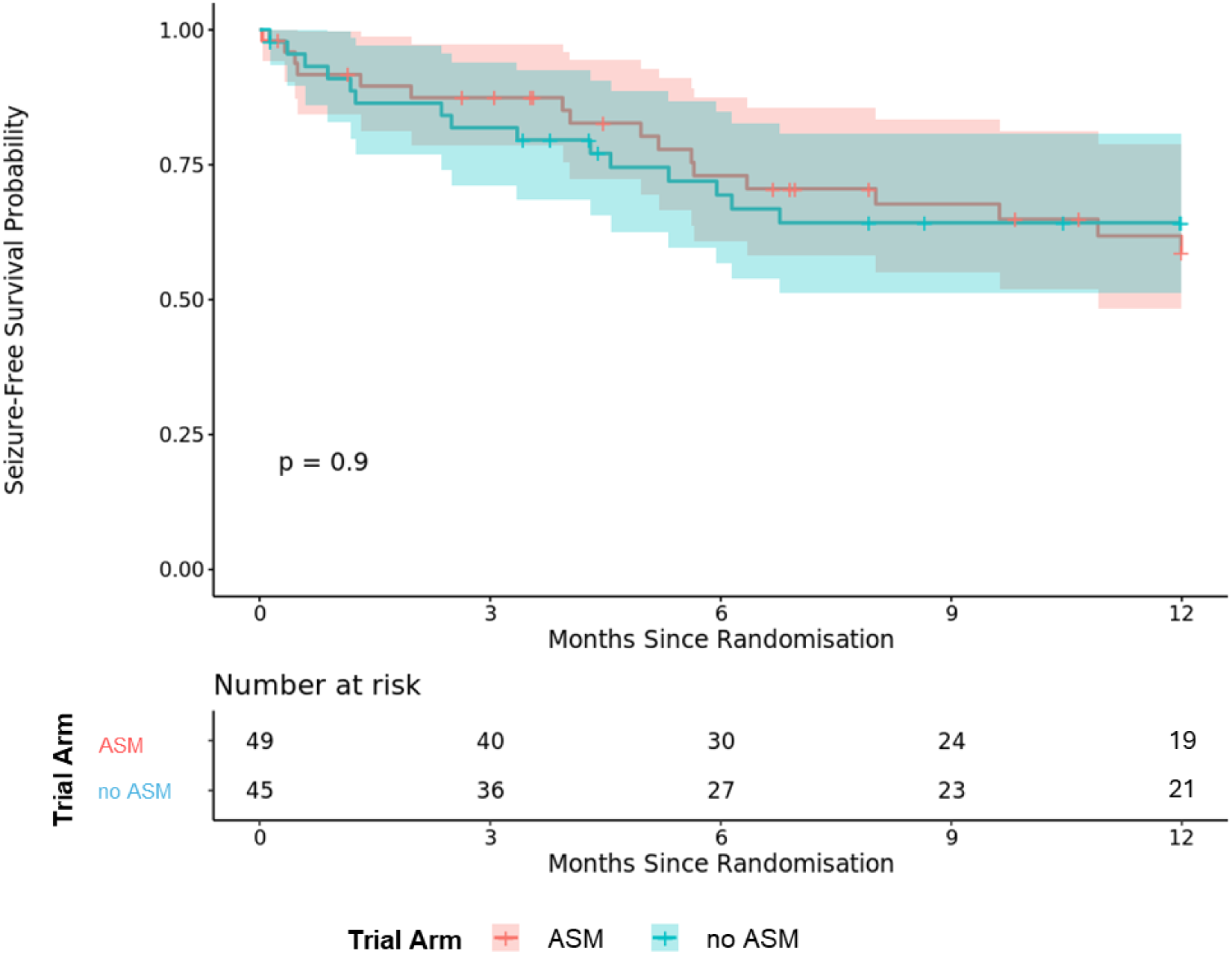
Kaplan Meier curve showing time to first seizure for the whole study population. The numbers of patients at risk for each group is shown. The p-value for the log-rank test is greater than 0.05 (p=0.9), therefore the null hypothesis that there is no difference between groups in terms of the distribution of time until seizure can be rejected at the 5% level.

Thirteen patients had a tonic-clonic seizure and the median time to first tonic clonic seizure was 6.2-months in the levetiracetam group and 1.2-months in the no prophylaxis group (Table 2). There were insufficient numbers to perform a statistical analysis between the two groups.

**Table 2:** Number of patients who had any type of seizure or a tonic clinic seizure within one-year of randomization according to trial arm.

| **Trial Arm** | **Patients** | **Any seizure**<br>**(%)** | **Tonic clonic**<br>**seizure (%)** | **Median time to first**<br>**tonic clonic seizure**<br>**(months)** |
| --- | --- | --- | --- | --- |
| Intervention arm<br>(levetiracetam) | 49 | 17 (34.7) | 8 (16.3) | 6.2 |
| Control arm (no<br>prophylaxis) | 45 | 15 (37.5) | 5 (11.1) | 1.2 |
| **Total** | **94** | **32 (34)** | **13 (13.8)** | **4.6** |

Progression free survival was not statistically different between the levetiracetam group (8.2-months) and the no prophylaxis group (10.4-months) (Figure S1). The 12-month overall survival rate was 59.2% in the levetiracetam group and 68.9% in the no prophylaxis group and the difference was not statistically significant (Figure S2). In the levetiracetam group 20 (41%) of 49 patients died within 12-months (median overall survival 6.8-months; range 0.2-11.9), and in the no prophylaxis group 14 (31%) of 45 patients died within 12-months (median 4.6-months; range 0.1-12.0).

The Semantic Verbal Fluency Test (SVLT: the number of animals patients can name in one minute) is a quick repeatable method to measure executive function, language and processing speed.(27) At baseline the median semantic verbal fluency test score was 18 (range: 3-40) in the levetiracetam arm, and 18.5 (range 2-30) in the no prophylaxis arm (Table S7). Completion rates were 97.8% at baseline and progressively decreased during the trial to 40.4% at 12-months. The median scores ranged between 17-18 in the levetiracetam group and 16-22 in the no prophylaxis group (Table S7). Anterograde memory test scores showed a similar decline in completion rates and scores were similar in both trial arms and remained static over 12-months (Figure S3). The Patient Health Questionnaire-9 (PHQ-9) is an instrument for screening and monitoring depressive symptoms. The percentage completion rates decreased at later points in the trial. Most patients reported PHQ-9 scores below 15, consistent with relatively mild depressive symptoms (Figure S4). Fatigue scores derived from the PHQ-9 Item 4 were similar in both groups at baseline, 3, 6, 9 and 12-months (Table S8). The Liverpool Impact of Epilepsy Scale (LIES)(6) and the Liverpool Seizure Severity Score (LSSS)(3) measure the impact and severity of epilepsy on different aspects of a person’s life. LIES scores ranged from 41 to 72 in the levetiracetam arm and 38 to 72 in the no prophylaxis arm (Figure S5). LSSS scores ranged from 34 to 44 in the levetiracetam arm and 35 to 49 in the no prophylaxis arm (Figure S6).

The mean EQ-5D-5L utility scores by time point and intervention arm (Table S9) were similar across time points and intervention arm. At baseline and 3-months the mean utility values were slightly higher in the no prophylaxis group (0.807 and 0.768) as compared with the levetiracetam group (0.780 and 0.747). At 6-months and 9-months the mean utility values were slightly higher in the levetiracetam group (0.786 and 0.759) as compared with the no prophylaxis group (0.748 and 0.723). At 12-months, the mean utility value in the no prophylaxis group (0.804) was higher as compared with the levetiracetam group (0.733). Mean QALYs by time point and intervention arm are shown in (Table S10). The mean number of QALYs accrued is very similar between the no prophylaxis group and levetiracetam group at each time point.

The mean cost of the intervention cost per person was estimated to be between £231 and £254 per 3-month period (Table S11). At baseline the mean total health care costs in the previous 3-months were very similar (£2580 in the levetiracetam group and £2532 in the no prophylaxis group). At both the 3-month and 6-month data collection points the total health care costs were lower in the levetiracetam group (£1175 and £1278) compared the no prophylaxis group (£2703 and £2767). At the 9-month and 12-month data collection points the total mean health care costs were higher in the levetiracetam group (£1916 and £1238) compared to the no prophylaxis group (£1597 and £686) (Table S12). Given the small sample sizes, no firm conclusions can be made from either the HRQoL or cost data.

In the patients taking levetiracetam, seven reported severe depression and/or suicidal ideation, of which five stopped the medication and withdrew from the trial and two stopped the medication but stayed on trial. The median time to stopping levetiracetam was 196 days (range: 18-295 days). Two patients had a CTCAE grade 2 adverse reaction (somnolence requiring levetiracetam dose reduction and altered mood and depression leading to patient withdrawal). There were 13 Serious Adverse Events reported that were unrelated to the levetiracetam. There were no SUSARs (Table S13).

## Discussion

In this randomised controlled trial of ASM prophylaxis in seizure-naïve patients with glioma undergoing surgery, the 12-month seizure rate was 47% in those receiving 12-months prophylactic levetiracetam, and 41% in those not receiving anti-seizure medication. The study closed early due to slow accrual as a consequence of the COVID pandemic, with only 94 of the planned 804 patients recruited and was underpowered to answer the primary research question. The study hypothesis was that prophylactic levetiracetam would increase the time to first seizure and reduce the severity of the first seizure should it occur. Time to first seizure was longer in the levetiracetam arm (5.0-months) compared to the no prophylaxis arm (2.5-months), and time to first tonic clonic seizure (a measure of seizure severity) was longer in the levetiracetam group (6.0-months) compared to the no prophylaxis arm (1.2-months). Whilst these findings suggest a direction of effect favouring the use of prophylactic levetiracetam the difference was not statistically significant, and as such does not support a change in routine clinical practice.

The SPRING trial is the largest randomised study to date on this question and provides additional data to inform the debate on the use of seizure prophylaxis in glioma surgery, although it does not fundamentally alter the current knowledge base. Current EANO/SNO clinical practice guidelines do not recommend the use of seizure prophylaxis for patients with newly diagnosed brain tumors (primary and metastatic).(26) These guidelines systematically reviewed the literature and rated the studies as class I (RCT), class II (cohort studies), or class III (case control studies). There were three RCT (class I), eight class II and eleven class III studies suggesting that seizure prophylaxis is not effective.(3) Only one class II study in melanoma metastasis (10) and one class III study in glioblastoma (18) supported their use. The recommendations are based mainly on lower quality studies, across a range or brain tumor types (primary and metastatic), and predominantly using first generation anti-seizure medication such as phenytoin and valproate. The SPRING trial, although underpowered, adds further support to the recommendation not to use seizure prophylaxis.

Up to one in four patients with brain tumors have side effects from anti-seizure medication that warrant a change or discontinuation.(9) Compared to first generation ASM, levetiracetam has a more favourable adverse effect profile (23) but is still associated with fatigue (15%), behavioural problems (13-38%) and aggression.(19) All these symptoms can also arise from the glioma, surgery, radiotherapy and chemotherapy. Whilst there is some evidence that levetiracetam may worsen these symptoms,(28) other studies have suggested that levetiracetam may be neuro-protective in brain injury (16) and that it may be associated with improved cognition in brain tumor patients.(7) In the SPRING trial there was no difference in mood, personality, fatigue and memory at any timepoint between the two trial arms. These data should be interpreted with caution, due to the low number of patients, and the inevitable attrition in data completeness at the 6-, 9- and 12-month timepoints. It should be noted that seven patients in the levetiracetam arm withdrew from the trial or stopped the medication due to severe depression or suicidal ideation. Consequently, in addition to their being no difference in seizure rates, time to first seizure and seizure severity, levetiracetam may be causing harm with no benefit.

Levetiracetam may have an oncological benefit for patients taking the alkylating chemotherapy drug temozolomide.(4) Poor quality retrospective studies, with significant risk of bias, have reported increased progression free and overall survival in patients with glioblastoma who received levetiracetam prophylactically or for seizures compared to those that didn’t(14 20 22) although data from a systematic review does not support this finding.(26) In the SPRING trial, there was no difference in progression free or overall survival between the two treatment arms. A trial to determine the efficacy of levetiracetam on survival in glioblastoma would likely face similar recruitment challenges.(14)

Seizures have an impact on quality of life and cause uncertainty and anxiety for patients and carers.(2) Seizures have an associated healthcare cost due to unscheduled admission to emergency departments. In the SPRING trial, there was a lower mean level of health care costs in the levetiracetam group compared with the no prophylaxis group at 3 and 6-months, however, at 9 and 12-months, mean health care costs were higher in the levetiracetam group. Health related quality of life was similar in both arms across all time points. None of the data on costs or HRQoL should be taken as any evidence of differences in HRQoL and cost. A definitive clinical trial combined with long-term economic modelling is still needed to fully understand the economic implications of ASMs for patients undergoing glioma surgery.

The strengths of this study are that: (i) despite early closure this was the largest randomised controlled trial to date to investigate the role of prophylactic anti-seizure medication to date in patients undergoing glioma surgery; (ii) the study investigated prolong seizure prophylaxis over 12-months; (iii) the study included both low and high grade glioma and these were balanced across each arm; (iv) participants were recruited across the whole of the UK to encompass adults of all ages and socio-economic classes and there was no loss to follow-up; (v) the completion rates for the health economics measures (EQ-5D-5L and heath care utilisation questionnaire) were good, implying that the measures were fit for purpose and could be used in a future, definitive study.

Some limitations of the trial should be noted. First, it was not possible to blind the participants and researchers as this was an open-label trial. This was an intentional part of the design since it was considered more pragmatic to deliver and pre-trial work with patients, carers and charities showed that administering a placebo in patients with a limited life-expectancy was not acceptable, and that the primary outcome measure was less susceptible to a placebo effect. Second, approximately one-quarter of the participants died from their glioma within 12-months and therefore did not reach the primary outcome of seizure rate at 12-months. Finally, the study was underpowered and closed early due to slow recruitment impacted by the COVID-19 pandemic, and we cannot fully exclude a difference between the two arms that could favor prophylaxis.

In conclusion, the SPRING trial is the largest prospective randomized study of seizure prophylaxis in glioma surgery worldwide. The study closed early and suffered from poor accrual caused by the COVID-19 pandemic. Although the study is underpowered, based on the limited data there was no difference in the 12-month seizure risk in the prophylaxis and no prophylaxis arms. The wide confidence intervals mean that we cannot be certain that a difference exists. The study provides the highest quality data available that could be combined with other studies for a future individual patient data meta-analysis. Current national and international guidelines will continue advise against the use of seizure prophylaxis. A definitive clinical trial is still needed to answer this important clinical question.

## Data Availability

Individual participant data that underlie the results reported in this article, after deidentification, will be made available. The study protocol, statistical analysis plan and consent forms will also be made available. Data will be available beginning 9-months and ending 3 years after publication. Data will be available to researchers whose proposed use of the data is approved by the original study investigators. Proposals should be directed to the corresponding authors and requestors will need to sign a data access agreement.

## Authorship

<u>Chief investigators</u>: RG and later MDJ (upon RG retirement)

<u>Study design</u>: RG, MDJ, HB, AGM

<u>Study and protocol development</u>: RG, MDJ, HB, JB, AR, JC, SL, TR, LV, GT, SCE, CW, AGM

<u>Trial management group member</u>: RG, MDJ, HB, JB, AR, JC, SL, TR, LV, GT, SCE, CW, AGM

<u>Statistical analysis</u>: RD, JC, RW, CG

<u>Health economic analysis</u>: TR, LV

<u>Clinical interpretation</u>: RG, MDJ, SCE, GT, TR, LV, HB,

<u>Drafting, editing and final report approval</u>: All authors

## Disclaimer

The views expressed are those of the authors and not necessarily those of the NHS, the NIHR or the Department of Health and Social Care. UCB Pharma did not have any intellectual input or oversight in the conduct and reporting of the SPRING trial.

## Acknowledgements

We acknowledge the members of the trial steering committee and independent data monitoring committee (see supplementary appendix pp2).

