## Supplementary Appendix for "Seizure prophylaxis in glioma surgery (SPRING): a multi-centre, unblinded, randomised trial"

This appendix has been provided by the authors to give readers additional information about their work.

SPRING trial Supplementary Material – table of contents

1. Trial oversight committees and list of investigators and recruitment by center (pages 2-3)

2. Trial flowchart (page 4)

3. Additional tables and analyses (pages 5-17)

- Figure S1 –(page 5)
- Figure S2 –(page 6)
- Figure S3 –(page 7)
- Figure S4 (page 8)
- Figure S5 (page 9)
- Figure S6 (page 9)
- Table S1 –(page 10)
- Table S2 –(page 11)
- Table S3 –(page 12)
- Table S4 – (page 12)
- Table S5 –(page 12)
- Table S6 –(page 13)
- Table S7 –(page 13)
- Table S8 - (page 14)
- Table S9 – (page 14)
- Table S10 – (page 15)
- Table S11 - (page 15)
- Table S12 - (page 16)
- Table S13 - (page 17)

4. List of substantial amendments to SPRING protocol by version (pages 18-19)

### 1. Trial committees and list of investigators

**Trial Steering Committee (TSC) independent members:**
Professor Iain MacPherson (chair)
Dr John Paul Leach
Professor Martin van den Bent
Professor William Hollingworth
Dr Mark Gilbert
Miss Cat Graham
Dr Stuart Farrimond

**Independent Data Monitoring Committee (IDMC) members:**
Professor Peter Langhorne (chair)
Professor Robert Hills
Dr Carl Counsell

***Trial writing group***
The writing group members are listed in the main author byline. MDJ and RG are the first and senior authors respectively and along with HB, AGR, JB and RD wrote the first draft of the manuscript, which was revised and approved by all the authors, who also assume responsibility for the accuracy and completeness of its content. The decision to submit the manuscript for publication lies with MDJ and RG. Statistical analysis was performed by RD, JC, RW, and CG according to the statistical analysis plan.

**Participating Sites and investigators and recruitment by centre**

| Centre / Hospital (number recruited) | PI | Co-investigators |
|---|---|---|
| Walton Centre Liverpool (18) | Michael D Jenkinson (CI) | Farouk Olubjo<br>Andrew R Brodbelt<br>David DA Lawson<br>Rasheed Zakaria<br>Emmanuel Chavredakis<br>Deborah Davies<br>Sikhangezile Gwatikunda<br>Samantha J Mills |
| Western General Hospital Edinburgh (16) | Imran Liaquat | Robin Grant (CI)<br>Paul Brennan<br>Sharon Peoples<br>David Summers<br>Kirsty Peebles |
| Kings College Hospital (12) | Keyoumars Ashkan | Katia Cikurel<br>Maria Alexandra Velicu<br>Natalie Long |
| Hull Royal Infirmary (12) | Chittoor Rajaraman | Shailendra Achawal<br>Adam Razak<br>Sanjay Dixit<br>Hiten Joshi<br>Emma Clarkson |
| Southampton General Hospital (8) | Paul Grundy | Harriet Joy<br>Jeng Ching<br>Mirriam Taylor |

| | | |
|---|---|---|
| Royal Preston Hospital (8) | Isaac Phang | Erica Beaumont<br>Sachin Mathur<br>Allan Brown |
| Leeds General Infirmary (7) | Ryan Mathew | Elizabeth Culpin<br>Mary Kambafwile |
| Addenbrookes Hospital (7) | Stephen J Price | Rajesh Jena<br>Tomasz Matys<br>Lynn Cream |
| Salford Royal Hospital (4) | Pietro D'Urso | Rovel Colaco<br>Ibrahim Djoukhadar<br>Kandeel Batool |
| Royal Stoke Hospital (3) | Erminia Albanese | Sumera Butt<br>Jooly Joseph<br>Ida Ponce |
| Queen Elizabeth Birmingham (1) | Shanika Samarasekera | Colin Watt<br>Victoria Wykes<br>Helen Benghiat<br>Satheesh Ramalingam |
| Queen Elizabeth Glasgow (0) | Athanasios Grivas | |
| Charing Cross Hospital (0) | Matthew Williams | |
| John Radcliffe Hospital (0) | Puneet Plaha | |

### 2. Trial flowchart

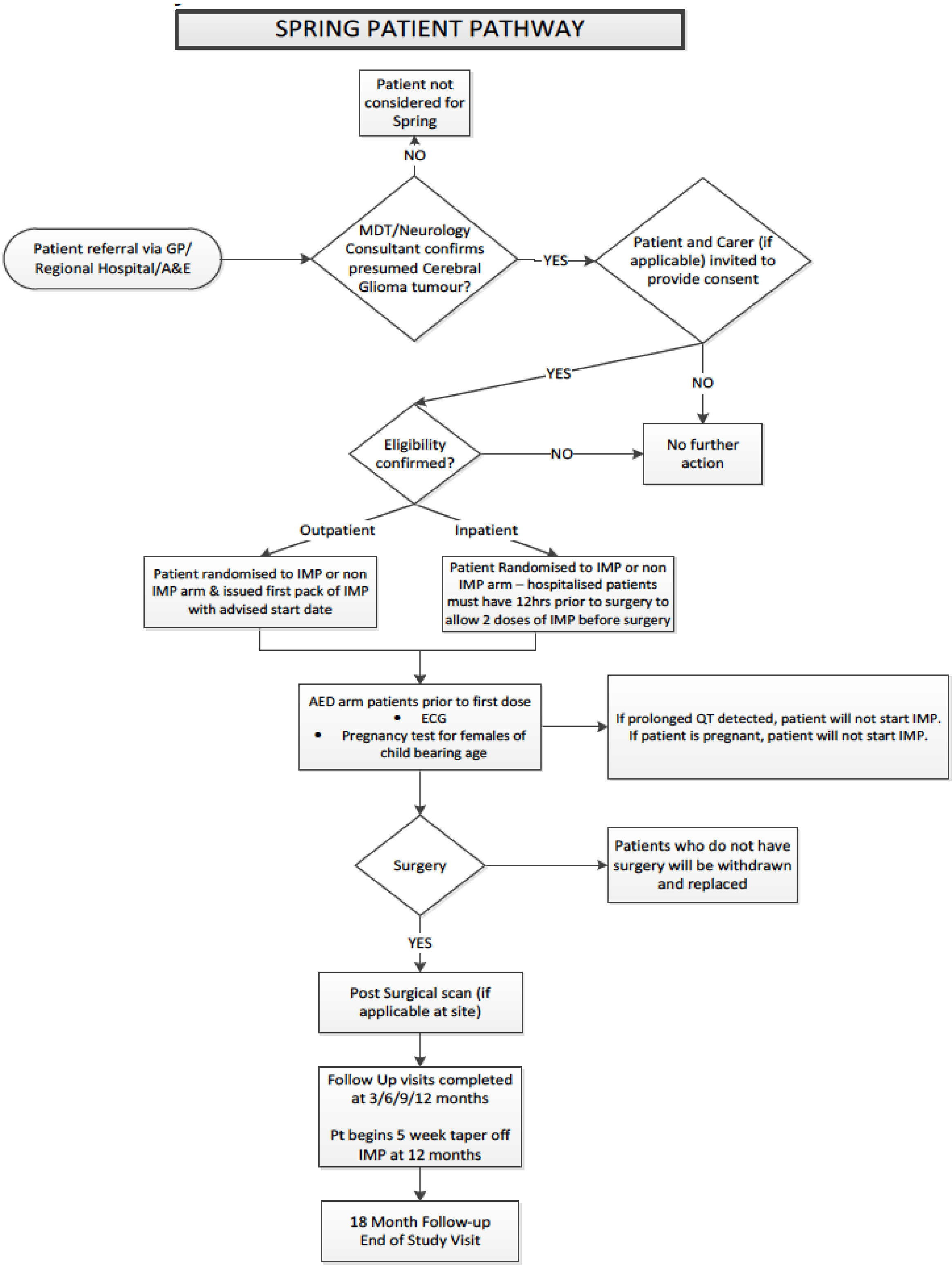


MDT: multi-disciplinary team
AED: Anti-epileptic drug
IMP: investigational medicinal product

**3. Additional figures, tables and analyses**

***Supplementary Figure S1***

Kaplan Meier curve showing time to progression. The p-value for the log-rank test is greater than 0.05 (p=0.23), therefore the null hypothesis that there is no difference between groups in terms of the distribution of time until progression cannot be rejected at the 5% level.

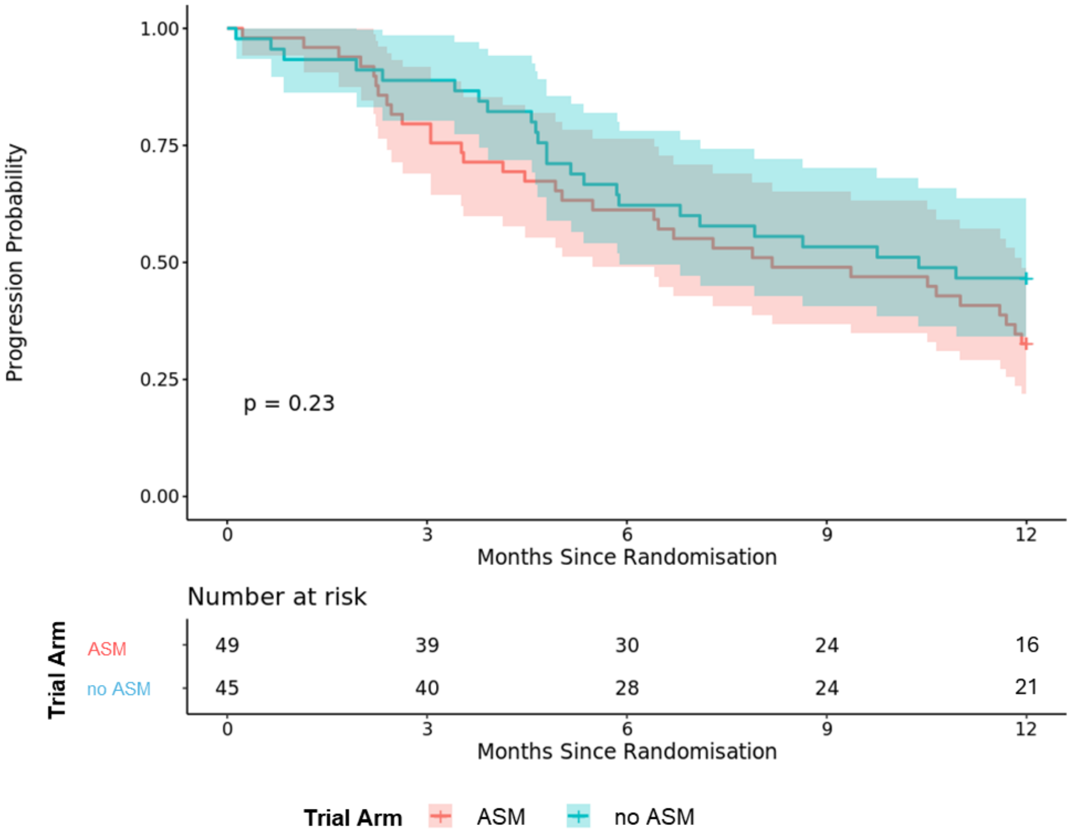

***Supplementary Figure S2***

Kaplan Meier curve showing time to death. The p-value for the log-rank test is greater than 0.05 (p=0.36), therefore the null hypothesis that there is no difference between groups in terms of the distribution of time until death cannot be rejected at the 5% level.

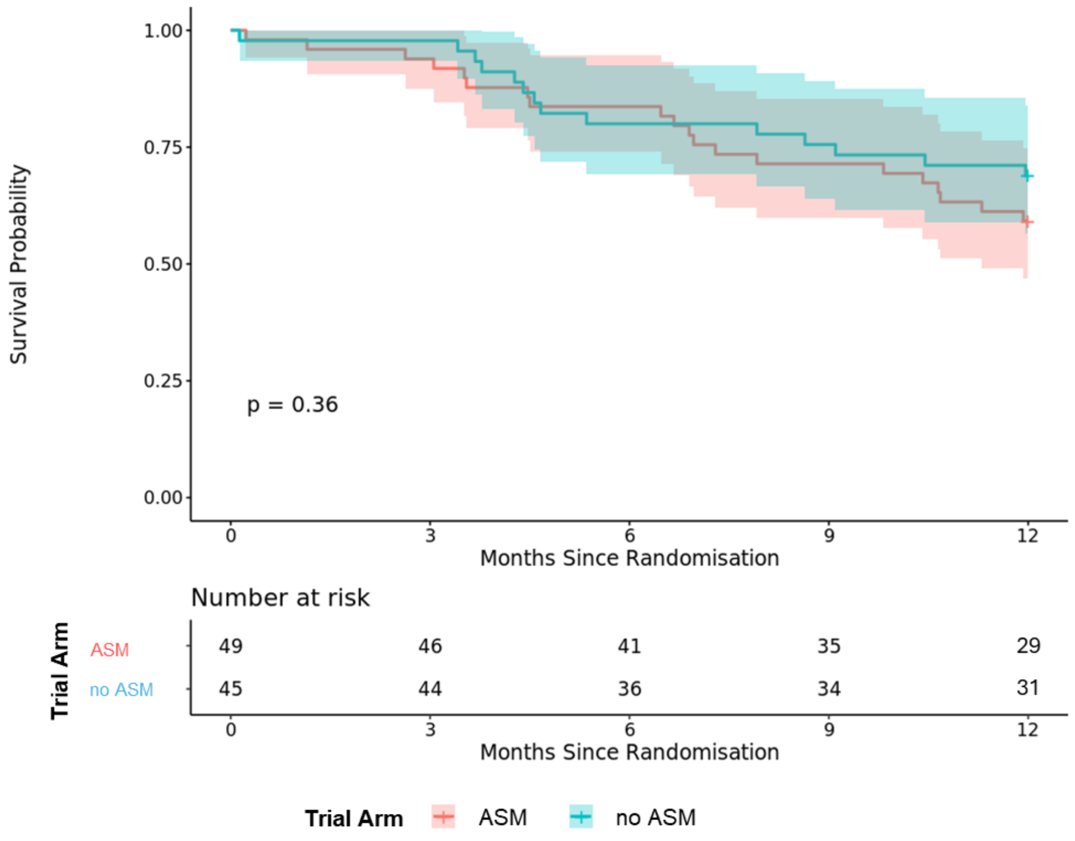

***Supplementary Figure S3***

Box plots showing the anterograde memory test score at baseline, 3, 6, 9 and 12 months.

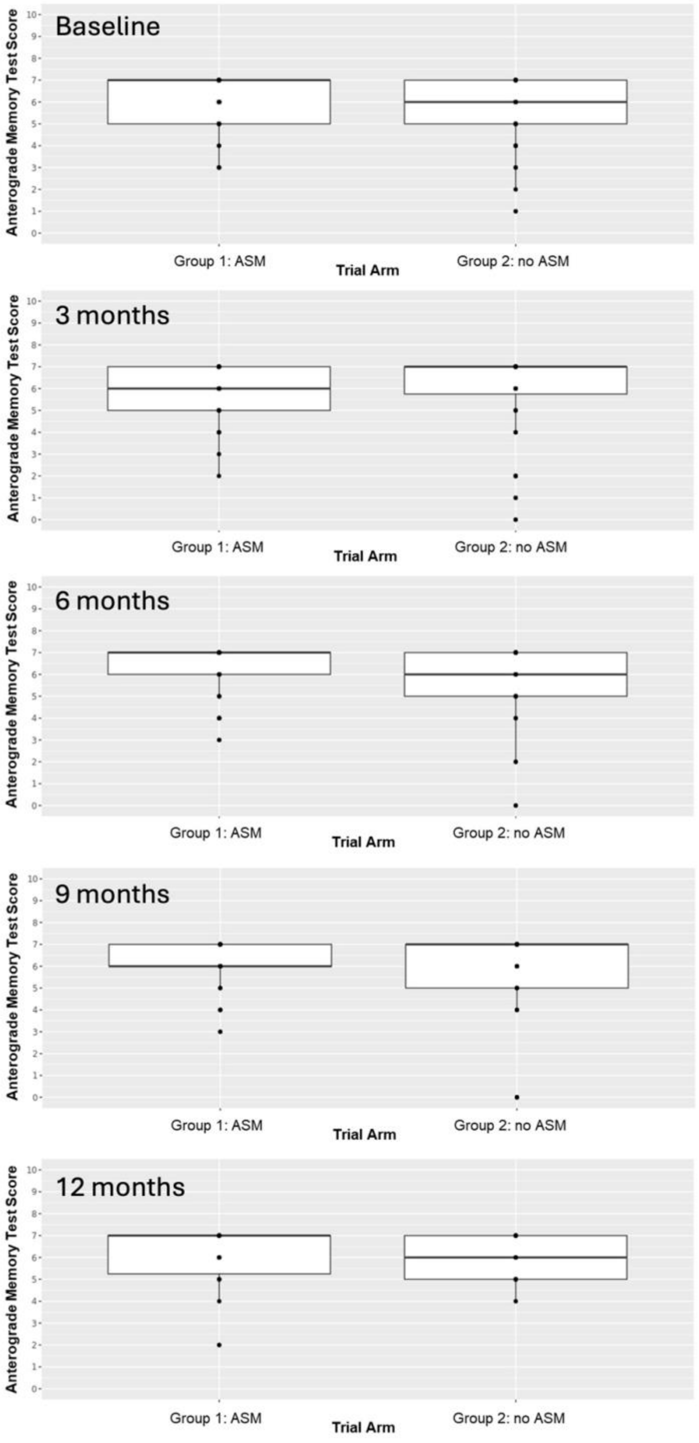

***Supplementary Figure S4***

Histogram plots showing the distribution of overall PHQ-9 test score at baseline, 3, 6, 9 and 12 months for the levetiracetam (pink) and no prophylaxis (green) groups.

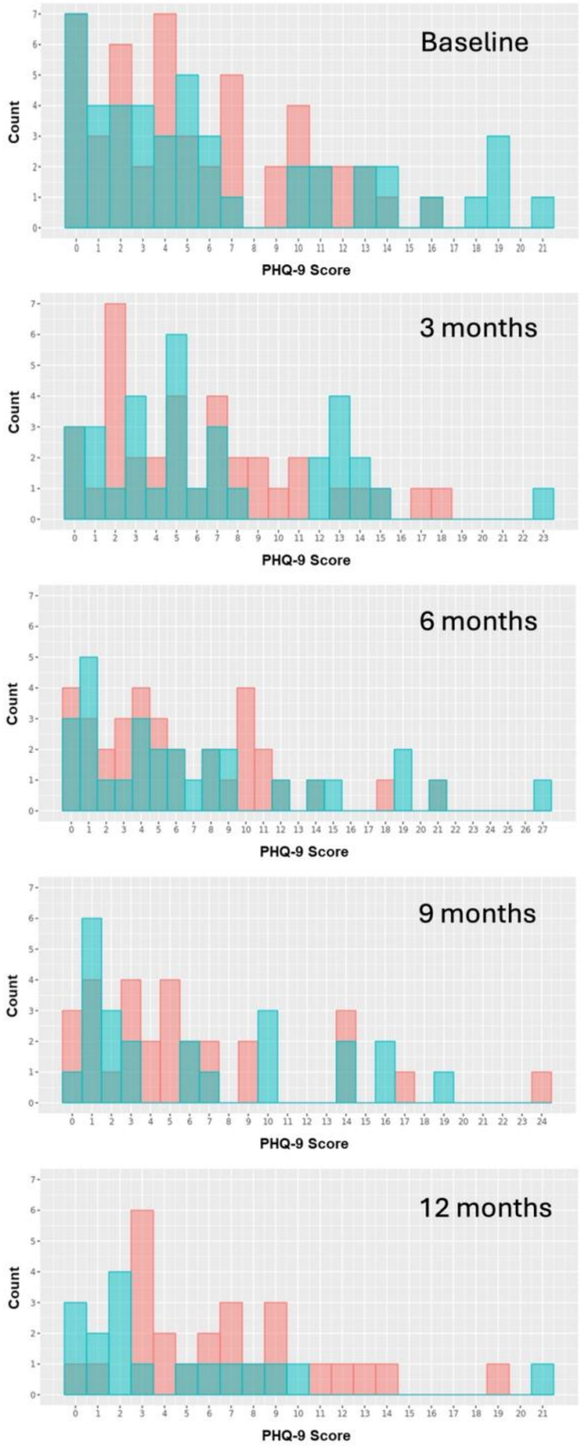

***Supplementary Figure S5***

Boxplots showing the distribution of tests scores for the Liverpool Impact of Epilepsy Scale. Higher scores are associated with a greater negative impact on daily quality of life.

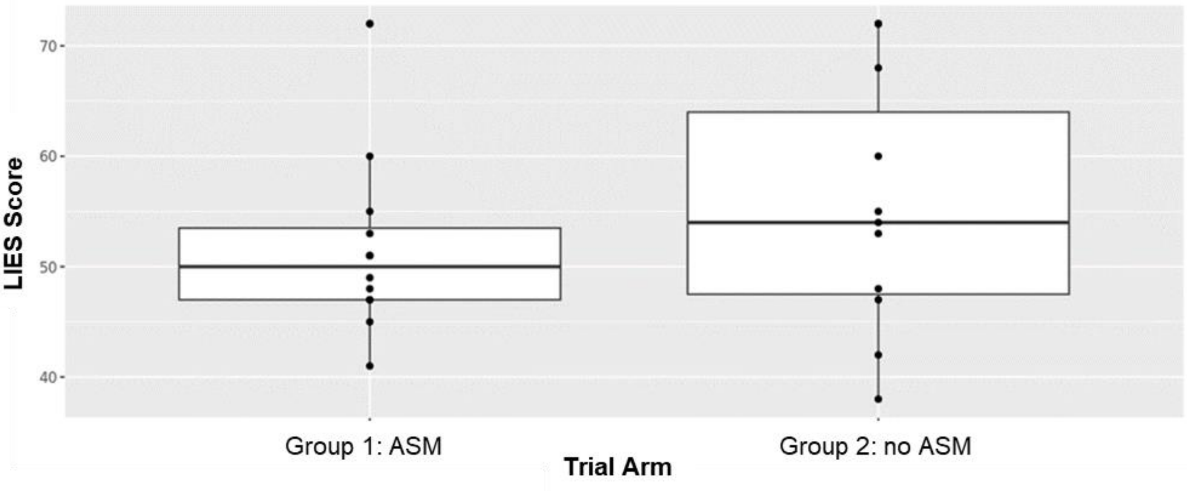


***Supplementary Figure S6***

Boxplots showing the distribution of tests scores between the levetiracetam (ASM) and no prophylaxis (no ASM) groups for the Liverpool Seizure Severity Score. Higher scores are associated with a greater negative impact on daily quality of life.

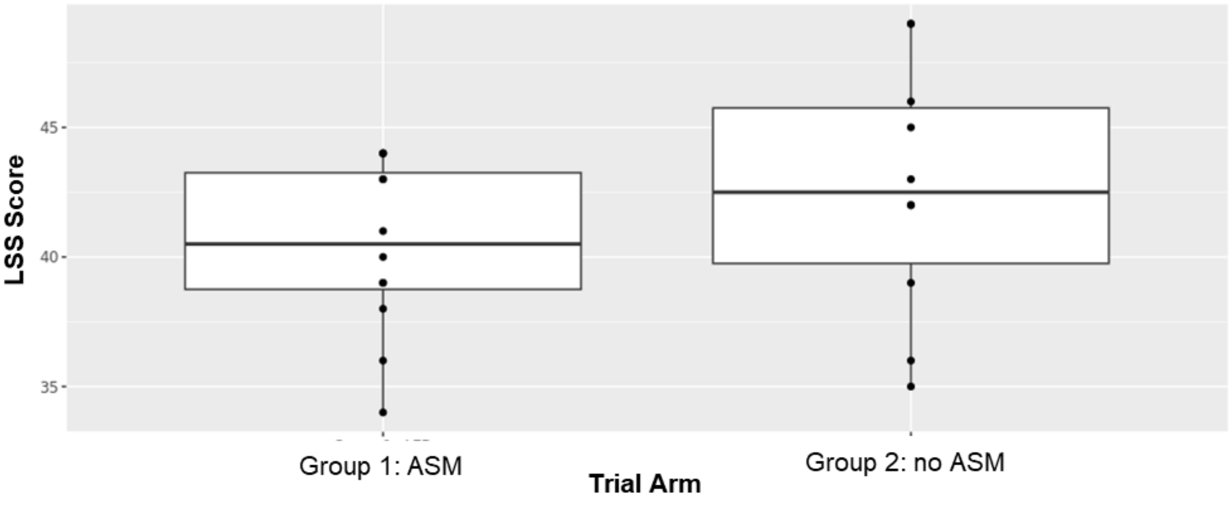

***Supplementary Table S1***

Levetiracetam dose titration and tapering schedule

| Normal treatment | Impaired Renal Function treatment |
|---|---|
| **Randomisation-2 weeks**<br>500 mg AM<br>500 mg PM | **Randomisation-2 weeks**<br>250 mg AM<br>250 mg PM |
| **Next 12-months**<br>750 mg AM<br>750 mg PM | **Next 12-months**<br>500 mg AM<br>500 mg PM |
| ***Week 1***<br>1 tablet of 500 mg + 1 tablet of 250 mg = 750mg AM<br>1 tablet of 500 mg = 500 mg PM | ***Week 1***<br>1 tablet of 500 mg = 500 mg AM<br>1 tablet of 250 mg = 250 mg PM |
| ***Week 2***<br>1 tablet of 500 mg = 500 mg AM<br>1 tablet of 500 mg = 500 mg PM | ***Week 2***<br>1 tablet of 250 mg = 250 mg AM<br>1 tablet of 250 mg = 250 mg PM |
| ***Week 3***<br>1 tablet of 500 mg = 500 mg AM<br>1 tablet of 250 mg = 250 mg PM | ***Week 3***<br>1 tablet of 250 mg = 250 mg AM |
| ***Week 4***<br>1 tablet of 250 mg = 250 mg AM<br>1 tablet of 250 mg = 250 mg PM | *No further treatment* |
| ***Week 5***<br>1 tablet of 250 mg = 250 mg AM | *No further treatment* |
| ***Week 6***<br>*No further treatment* | *No further treatment* |

***Supplementary Table S2***

Patient visit schedule

| | **Screening / pre-surgery** | **Post-surgery** | **3 month (+/- 2 wks)** | **6, 9, 12 month (+/- 2 wks)** | **18 month [a] (+/- 2 wks)** | **Reported suspected seizure** |
|---|---|---|---|---|---|---|
| Written informed consent | X | | | | | |
| Demographic data | X | | | | | |
| Medical /seizure history | X | | X | X | | |
| Concomitant medications | X | | X | X | **S** - details of AEDs only | |
| PHQ9 | X | | X | X | | |
| Karnofsky Performance Status (Neurological exam) | X | | X | X | | |
| Semantic Verbal Fluency Test (SVFT) (Neurological exam) | X | | X | X | | |
| Anterograde Memory test (Neurological exam) | X | | X | X | | |
| Tissue | **S** | | | | | |
| Blood (eGFR -kidney function) | **S** | | **S** | **S** | | |
| ECG[d] | X | | | | | |
| Pregnancy testing[c] | X | | | | | |
| Standard MRI imaging | **S** | **S** | <u>**S**</u> | | | |
| Liverpool Adverse Events Profile | X | | X | X | | |
| EQ-5D-5L | X | | X | X | | X |
| Carer EQ-5D-5L | X | | X | X | | |
| Liverpool Impact of Epilepsy Scale | | | | | | X |
| Liverpool Seizure Severity Scale | | | | | | X |
| Health Economic Questionnaire | X | | X | X | | |
| Time & Travel Questionnaire[b] | | | | X | | |

*Key:* X – Completed as Trial activity **S –** Standard of care, to be completed as per institution guidelines <u>**S -**</u> Standard of care, to be completed as per institution guidelines, 3 month MRI will capture institution MRI between post-surgery and up to 6 months.

[a] *Follow up will be every 3 months to at least one year after randomisation, where possible 18 month data will be collected until the end of trial* [b] *Time and Travel Questionnaire is at 6 month visit only* [c] *Pregnancy test should take place before first dose of IMP (if on IMP arm)* [d] *ECG before first dose of IMP (if on IMP arm)*

***Supplementary Table S3***
Karnofsky Performance Status for each trial arm

| **Karnofsky Performance Status** | **Intervention arm (Levetiracetam)** | **Control arm (no prophylaxis)** | **Total** |
|---|---|---|---|
| Normal no complaints; no evidence of disease | 16 | 11 | 27 |
| Able to carry on normal activity; minor signs or symptoms of disease. | 18 | 22 | 40 |
| Normal activity with effort; some signs or symptoms of disease. | 5 | 4 | 9 |
| Cares for self; unable to carry on normal activity or to do active work. | 10 | 8 | 18 |
| Requires occasional assistance, but is able to care for most of his personal needs. | 0 | 0 | 0 |
| Missing | 0 | 0 | 0 |
| **Total** | **49** | **45** | **94** |

***Supplementary Table S4***
Anatomical location of gliomas based on the pre-surgery MRI. Gliomas can be location in multiple anatomical areas

| **Tumour Location*** | **Intervention arm (Levetiracetam)** | **Control arm (no prophylaxis)** | **Total** |
|---|---|---|---|
| Basal Ganglia | 1 | 2 | 3 |
| Cerebellum | 1 | 0 | 1 |
| Corpus Callosum | 7 | 5 | 12 |
| Frontal | 21 | 19 | 40 |
| Insular | 8 | 6 | 14 |
| Occipital | 4 | 13 | 17 |
| Parietal | 13 | 13 | 26 |
| Brainstem | 0 | 1 | 1 |
| Temporal | 14 | 21 | 35 |
| Thalamus | 3 | 2 | 5 |
| *Not mutually exclusive | | | |

***Supplementary Table S5***
Histopathology diagnosis according to the WHO Classification of Tumours.

| **Glioma Histology** | **Intervention arm (Levetiracetam)** | **Control arm (no prophylaxis)** | **Total** |
|---|---|---|---|
| Diffuse Astrocytoma (Grade 2) | 1 | 1 | 2 |
| Anaplastic Astrocytoma (Grade 3) | 1 | 1 | 2 |
| Glioblastoma (Grade 4) | 42 | 37 | 79 |
| Other Astrocytoma | 2 | 2 | 4 |
| Oligodendroglioma | 0 | 3 | 3 |
| Anaplastic Oligodendroglioma | 0 | 0 | 0 |
| Other Oligodendroglioma | 0 | 0 | 0 |
| Other Glioma | 3 | 1 | 4 |
| Missing | 0 | 0 | 0 |
| **Total** | **49** | **45** | **94** |

***Supplementary Table S6***

Molecular features of the glioma according to treatment arm.

| Genotype Marker | Intervention arm (Levetiracetam) | Control arm (no prophylaxis) | Total |
|---|---|---|---|
| **IDH2** | | | |
| IDH mutant | 5 | 11 | 16 |
| IDH wild type | 39 | 33 | 72 |
| Not done | 4 | 1 | 5 |
| Missing | 1 | 0 | 1 |
| Total | 49 | 45 | 94 |
| **MGMT** | | | |
| Methylated | 21 | 18 | 39 |
| Unmethylated | 24 | 19 | 43 |
| Not done | 3 | 8 | 11 |
| Missing | 1 | 0 | 1 |
| Total | 49 | 45 | 94 |
| **Chromosome 1p** | | | |
| Co-deleted | 2 | 5 | 7 |
| Non co-deleted | 12 | 10 | 22 |
| Not done | 33 | 29 | 62 |
| Missing | 2 | 1 | 3 |
| Total | 49 | 45 | 94 |
| **TERT** | | | |
| Mutated | 10 | 14 | 24 |
| Wild type | 2 | 2 | 4 |
| Not done | 35 | 27 | 62 |
| Missing | 2 | 2 | 4 |
| Total | 49 | 45 | 94 |

***Supplementary Table S7***

Semantic Verbal Fluency Test (SVLT) scores for the levetiracetam and no prophylaxis arms at baseline, 3, 6, 9 and 12 months.

| Trial Arm | N | Median | Range | Standard Deviation | Missing (%) |
|---|---|---|---|---|---|
| **Baseline** | | | | | |
| Levetiracetam (Intervention arm) | 49 | 18.0 | 3-40 | 7.5 | 0 (0) |
| No prophylaxis (Control arm) | 45 | 18.5 | 2-30 | 7.6 | 1 (2.2) |
| **3 months** | | | | | |
| Levetiracetam (Intervention arm) | 49 | 18 | 3-35 | 7.4 | 14 (28.6) |
| No prophylaxis (Control arm) | 45 | 16 | 0-35 | 8.3 | 9 (20.0) |
| **6 months** | | | | | |
| Levetiracetam (Intervention arm) | 49 | 18 | 3-27 | 6.9 | 17 (34.7) |
| No prophylaxis (Control arm) | 45 | 17 | 0-41 | 9.4 | 17 (37.8) |
| **9 months** | | | | | |
| Levetiracetam (Intervention arm) | 49 | 18.0 | 5-29 | 5.5 | 22 (44.9) |
| No prophylaxis (Control arm) | 45 | 19.5 | 1-49 | 11.2 | 21 (46.7) |
| **12 months** | | | | | |
| Levetiracetam (Intervention arm) | 49 | 17 | 4-27 | 5.8 | 27 (55.1) |
| No prophylaxis (Control arm) | 45 | 22 | 7-50 | 12.1 | 29 (64.4) |

***Supplementary Table S8***

Fatigue scores from the PHQ-9 test for the levetiracetam and no prophylaxis arms at baseline, 3, 6, 9 and 12 months. Patients report how often they have been 'feeling tired or having little energy' over the preceding 2 weeks. Patients score as: 0=not at all, 1=several days, 2=more than half the days, and 3=nearly every day.

| Trial Arm | N | Median | Range | Missing (%) |
|---|---|---|---|---|
| **Baseline** | | | | |
| Intervention arm (Levetiracetam) | 49 | 1 | 0-3 | 0 (0) |
| Control arm (no prophylaxis) | 45 | 1 | 0-3 | 0 (0) |
| **3 months** | | | | |
| Intervention arm (Levetiracetam) | 49 | 1 | 0-3 | 13 (26.5) |
| Control arm (no prophylaxis) | 45 | 1 | 0-3 | 12 (26.7) |
| **6 months** | | | | |
| Intervention arm (Levetiracetam) | 49 | 1 | 0-3 | 15 (30.6) |
| Control arm (no prophylaxis) | 45 | 2 | 0-3 | 16 (35.6) |
| **9 months** | | | | |
| Intervention arm (Levetiracetam) | 49 | 1 | 0-3 | 20 (40.8) |
| Control arm (no prophylaxis) | 45 | 1 | 0-3 | 22 (48.9) |
| **12 months** | | | | |
| Intervention arm (Levetiracetam) | 49 | 1 | 0-3 | 24 (49.0) |
| Control arm (no prophylaxis) | 45 | 1 | 0-3 | 28 (62.2) |

***Supplementary Table S9***

Mean EQ-5D-5L Utility Score by Intervention Arm (Self)

| **Self-Completed** | |
|---|---|
| **Intervention Arm (ASM)** | **Control Arm (No ASM)** |
| n = 48 | n = 43 |
| 0.780 (0.197) | 0.807 (0.185) |
| n = 30 | n = 30 |
| 0.747 (0.296) | 0.768 (0.254) |
| n = 31 | n = 26 |
| 0.786 (0.216) | 0.748 (0.271) |
| n = 28 | n = 23 |
| 0.759 (0.214) | 0.723 (0.320) |
| n = 23 | n = 17 |
| 0.733 (0.300) | 0.804 (0.167) |
| **Proxy-Completed** | |
| n = 25 | n = 28 |
| 0.751 (0.147) | 0.730 (0.193) |
| n = 19 | n = 23 |
| 0.696 (0.295) | 0.671 (0.293) |
| n = 20 | n = 19 |
| 0.673 (0.301) | 0.661 (0.278) |
| n = 17 | n = 16 |
| 0.698 (0.199) | 0.526 (0.422) |
| n = 15 | n = 7 |
| 0.654 (0.277) | 0.672 (0.229) |
| Mean values with standard deviations in parentheses. | |

***Supplementary Table S10***
Mean QALYs by Intervention Arm (Self – Completed)

| | **Intervention Arm (ASM)** | **Control Arm (No ASM)** |
|---|---|---|
| **3 Months** | n=42 | n=39 |
| | 0.162 (0.069) | 0.171 (0.059) |
| **6 Months** | n=42 | n=37 |
| | 0.295 (0.167) | 0.300 (0.154) |
| **9 Months** | n=41 | n=34 |
| | 0.416 (0.264) | 0.398 (0.245) |
| **12 Months** | n=39 | n=33 |
| | 0.501 (0.351) | 0.477 (0.331) |
| Mean values with standard deviations in parentheses. As no imputation methods were used for missing data, data is provided for complete cases only. The number of patients in each arm at each time point are different to those reported in Table A and Table B, as those patients who died during the trial period are assigned a utility value of 0 at all subsequent time points and therefore remain in the analysis sample. | | |

***Supplementary Table S11***
Mean Cost of Intervention (Levetiracetam) by Time Point

| **Mean Cost Per Patient** | 3 Months (n=40) | 6 Months (n=35) | 9 Months (n=30) | 12 Months (n=25) |
|---|---|---|---|---|
| Intervention Cost (£) | 254 (21) | 231 (20) | 233 (16) | 232 (17) |
| Notes: Mean values with standard deviations in parentheses. All values rounded to the nearest £. | | | | |

***Supplementary Table S12***
Mean Total Health Care Costs (£) by Time Point and Intervention Arm

| | **Intervention (ASM)** | | | | | **Control (No ASM)** | | | | |
|---|---|---|---|---|---|---|---|---|---|---|
| **Mean Cost Per Patient** | Baseline (n=49) | 3 Months (n=35) | 6 Months (n=34) | 9 Months (n=29) | 12 Months (n=24) | Baseline (n=44) | 3 Months (n=35) | 6 Months (n=30) | 9 Months (n=24) | 12 Months (n=17) |
| Primary Health Care | 169 (494) | 119 (398) | 155 (385) | 80 (101) | 78 (147) | 106 (100) | 92 (91) | 145 (188) | 152 (293) | 96 (106) |
| Secondary Health Care | 2360 (2410) | 912 (1558) | 942 (1083) | 1666 (5249) | 681 (654) | 2388 (2009) | 2531 (3043) | 2571 (4145) | 1208 (1396) | 520 (691) |
| Medication Use (Excluding Levetiracetam) | 51 (33) | 145 (484) | 181 (501) | 170 (377) | 480 (919) | 37 (34) | 80 (72) | 51 (49) | 237 (537) | 71 (188) |
| Total Health Care Cost | 2580 (2666) | 1175 (1709) | 1278 (1195) | 1916 (5299) | 1238 (1144) | 2532 (2022) | 2703 (3120) | 2767 (4169) | 1597 (1838) | 686 (724) |

Notes: Mean values with standard deviations in parentheses. All values rounded to the nearest £.

***Supplementary Table S13***
Serious Adverse Events reported in the patients taking levetiracetam. There were no SUSARs.

| Gender | Event Term | Event Outcome | Serious Criteria | Levetiracetam daily dose (mg) |
|---|---|---|---|---|
| Male | Neutropenic sepsis | Recovered | Hospitalisation | 750 |
| Male | Diarrhoea | Recovered | Hospitalisation | 1500 |
| Male | Cellulitis both legs | Recovered | Hospitalisation | 1500 |
| Female | Bowel perforation | Death | Death, Hospitalisation, Important medical event | 1500 |
| Male | Hospitalisation due to gall bladder infection | Recovered with sequelae | Hospitalisation | 1500 |
| Male | Provoked DVT | Recovered with sequelae | Hospitalisation | 1500 |
| Female | Pulmonary embolism | Recovered with sequelae | Hospitalisation | 1500 |
| Female | Cranial wound infection | Condition unchanged | Hospitalisation, Important medical event | 1500 |
| Male | Pneumonia | Recovered | Hospitalisation | 1500 |
| Female | Raised glucose levels | Recovered with sequelae | Hospitalisation | 1000 |
| Male | Respiratory distress | Death | Death, Hospitalisation | - |
| Female | Hypokalaemia | Recovered | Hospitalisation | 1500 |
| Male | Skin infection | Recovered | Hospitalisation | 1500 |

### 4. List of substantial amendments to SPRING protocol by version

| Protocol Version Number | Date Issued | Details of change(s) made |
|---|---|---|
| 6.0 | 15 May 2019 | • Reformatting of some sections within the protocol<br>• Addition of Section 1.5 (Disease Progression) to allow participants to stay in the study should they lose capacity after consenting<br>• More detailed description of LAEP and EQ-5D-5L (Section 2.5)<br>• Addition of PHQ9 questionnaire which is used as a screening tool for depressive symptoms. The protocol was also updated to remove patients with severe depression as defined by a PHQ-9 score of 20+<br>• Addition of Cognitive/capacity monitoring section (Section 2.6)<br>• Addition of Treatment Breaks section (Section 4.4) |
| 6.1 | 25 July 2019 | • Minor protocol amendment to clarify that if a patient takes another form of levetiracetam during a treatment break, this is not considered trial treatment |
| 6.2 | 25 September 2019 | • Minor updates to the protocol - Key contacts amended, section 5.3 (modified safety reporting clarified), section 7.3.5 (analysis performed for assessment of utility associated with seizures clarified), Appendix 3 (references updated) |
| 6.3 | 07 April 2020 | • Section 1.5 of the protocol (Side Effects of levetiracetam) updated to incorporate changes to section 4.8 of the SmPC - gait disturbance (walking disorder), encephalopathy, delirium, added as affecting between 1 in 1000 and 1 in 10000 people |
| 7.0 | 29 June 2020 | • Sponsor updated to Public Health Scotland<br>• Consent process updated to allow initial partial telephone consent for participants<br>• Remove pregnancy testing for IMP patients at follow up visits. As visits were 3 months apart, female participants of childbearing potential on the treatment arm advised to contact their site ASAP if they become pregnant. Female participants of childbearing potential on the treatment arm were asked at each visit if there was a possibility they have become pregnant, if they answered yes, a pregnancy test will be completed.<br>• Additional data collected for suspected COVID-19 illness dates and test results, following the COVID-19 pandemic.<br>• The PHQ-9 exclusion and withdrawal criteria amended from 20+ to ≥20, to improve clarity for sites |
| 8.0 | 24 May 2021 | • Section 1.5 of the protocol (Side Effects of levetiracetam) updated to incorporate changes to section 4.8 of the SmPC. "Seizures aggravated" and increased delay between heartbeats on Electrocardiogram" added as adverse drug reactions (ADR) affecting 1 in 1000 to 1 in 10000 people<br>• Patient pathway, (sections 3.4 and 4.3) updated to check for QT interval prolongation for patients randomised to the AED treatment arm before starting treatment. Any patient with QT interval ≥500 milliseconds will not start on IMP.<br>• Pregnancy testing updated in the patient pathway and sections 3.3, 3.4, 4.2, 4.3 and 5.7. Female patients of childbearing age randomised to the levetiracetam arm will have a pregnancy test to ensure they are not pregnant before starting trial medication. They will also complete a pregnancy test at any time during the trial if they confirm there is a possibility they could be pregnant.<br>• Sections 2.5 and 6.3 updated to introduce Electronic Patient Reported Outcomes (ePRO) giving participants and Carers the |

| | | |
|---|---|---|
| | | option to complete questionnaires online at home using REDCap.<br>• Section 7.3.5 (2) updated to introduce the Standard Gamble.<br>• Minor protocol amendments included to section 4.3 (MRI), 5.5 (SAEs), 2.5 and 4.2 (telephone EQ-5D-5L following first seizure) |
| 9.0 | 15 May 2023 | • Chief Investigator updated from Dr Robert Grant to Professor Michael Jenkinson<br>• Sponsor representatives amended |

The first protocol approved by the Research Ethics Committee was Version 5.1 on 04 February 2019
